# Knowledge and aptitude of early childhood, primary and/or secondary education teachers referred to first aid measures in dental trauma in the province of Seville (Spain.)

**DOI:** 10.1101/2022.01.18.22269338

**Authors:** David Ribas Pérez, Rosmery Olivera, Asunción Mendoza-Mendoza

## Abstract

The high of childhood dental trauma requires childcare professionals to have basic notions of dental first aid. The **aim** of this study is to assess the level of knowledge and aptitude (defined as the ability to operate competently in a given activity) of early childhood, primary and/or secondary education teachers from Seville (Spain) referred to first aid measures in dental trauma. A descriptive **study** was made. The study sample consisted of 442 teachers (334 women and 108 men) coinciding proportionately with the study target population in terms of gender distribution, type of center and teaching level.

**Methods:** A specifically designed questionnaire was used.

**Results:** Most of the participants (88.5%, n=391) exhibited poor knowledge and aptitude, 11.5% (n=51) showed medium level knowledge and aptitude, and none were categorized as having high knowledge and aptitude.

**Conclusion:** These findings could be explained by the fact that 98% of those surveyed had never received any training in this area.

## INTRODUCTION

The epidemiological data show the incidence of pediatric dental trauma (PDT) to be very high in dental practice [Lam, 2016]. Rapid and precise management is required in such situations, and close monitoring of the lesion is moreover required due to the possibility of early or late complications [Mendoza, 2003]. These indications are simple to follow in adult dental trauma, but prove more complex in the case of PDT, since such cases depend upon the intervention and knowledge of the patient caregiver at the time of the accident. Pediatric dental trauma has a number of consequences. Even when such injuries affect the temporary dentition, they may cause functional problems, affect the permanent tooth germ, produce infections and cause aesthetic problems with psychosocial consequences for the child. The degree of involvement is highly variable and depends on a range of factors [Lenzi et al., 2015; Mendoza et al., 2015; Diangelis et al., 2015.] All individuals in charge of childcare, particularly professionals, sports instructors and relatives, should be trained in what to do in situations of PDT. How to act at the site of the accident, and when and where to go, should be clear concepts with an established order, and should be applied correctly. It is crucial to invest in educational programs destined to ensure coherent action, since a few simple steps can improve the prognosis of dental trauma and may contribute enormously to the quality of our healthcare system [Pacheco et al., 2013; Young et al., 2013.]

Training requirements in pediatric caregivers In certain countries and sociocultural contexts, knowledge on the part of the general population regarding the origin of caries is acceptable, and there are studies indicating that an adult could mention at least three actions for preventing caries [Duijster et al., 2017.] International efforts in the form of global campaigns, as well as educational policies targeted to adults and children, are producing benefits [Hugoson et al., 2007.] Awareness-enhancing campaigns designed to promote tooth brushing are common in childhood centers [Angelopoulou et al., 2015.] In our setting (Andalusia, Spain), early childhood and primary and secondary education teachers have received training in this field and in other aspects such as fluoridation, thanks to the oral health promotion program “Aprende a Sonreír” (“Learn to Smile”) [Junta de Andalucia, 2020], which the regional health authorities have implemented since 2002. However, when it comes to dental trauma, these teachers evidence training shortcomings in many aspects, particularly as regards the provision of first aid at the site of the accident [Arikan and Sonmez, 2012.] Intervention in such situations on the part of the teaching staff and other childcare professionals directly conditions the prognosis of dental trauma. Knowledge among these individuals of first aid measures in dental trauma is therefore necessary [Diangelis et al., 2012; Andersson et al., 2017; Malmgrem et al. 2017.] Their actions are of crucial relevance particularly in certain clinical situations such as dental avulsión [Andersson et al., 2017.] Teachers should be aware that dental trauma is a clinical emergency [Mendoza 2003], and as such requires prompt attention from the pediatric dentist. They should be able to manage a clear and concise protocol with indications unifying professional criteria, and which are the same for all centers regardless of their public or private nature. Emphasis also must be placed on the need to prevent dental trauma through measures such as the use of mouth guards during sports activities [Ferrari and Ferreira De Medeiros, 2002; Persic et al., 2006.] A survey involving a representative sample of early childhood, primary, and/or secondary education teachers from the province of Seville (Spain) was carried out with the aim to evaluate their knowledge and aptitude (defined as the ability to operate competently in a given activity) referred to first aid measures in dental trauma.

## MATERIAL AND METHODS

A specific questionnaire was designed for our survey using Google Forms, with posterior validation by university lecturers specialized in the study topic. The collected data were entered on MS Excel spreadsheets for subsequent analysis using the SPSS statistical package. Study population and sample selection The study population of our survey comprised all active teachers during the school year 2019-2020 serving in public, private and/or concerted schools in the province of Seville (Spain), with teaching competences in early childhood, primary and/or secondary education (including high school and basic and middle level professional training). The health crisis caused by the COVID-19 pandemic made it necessary to administer the questionnaire online.

Sample size calculation for finite populations was carried out to determine the required number of completed questionnaires. Considering a population of 22,148 teaching professionals and assuming sample randomized sampling with a margin of error of 5%, the minimum number of questionnaires was seen to be 378 [Junta de Andalucía, 2019] (Table 1).

**Table 1.** Comparative distribution of the study population and sample in terms of gender, type of center and level of teaching.

|  |  | Population | Sample |
| --- | --- | --- | --- |
| **Gender** | Men | 27,47% | 24,40% |
|  | Women | 72,53% | 75,60% |
| **Type of Center** | Public | 69,28% | 76,50% |
|  | Private | 30,72% | 5,40% |
|  | Concerted |  | 18,10% |
| **Level of teaching** | Early Choldhood | 25,75% | 26,70% |
|  | Primary | 37,26% | 43,70% |
|  | Secondary | 36,99% | 29,60% |

### Study questionnaire

A specifically designed anonymous and self-administered questionnaire to be completed online was used for the study. The questionnaire was grouped into four groups with a total of 25 items (Table 2).

**Table 2.**
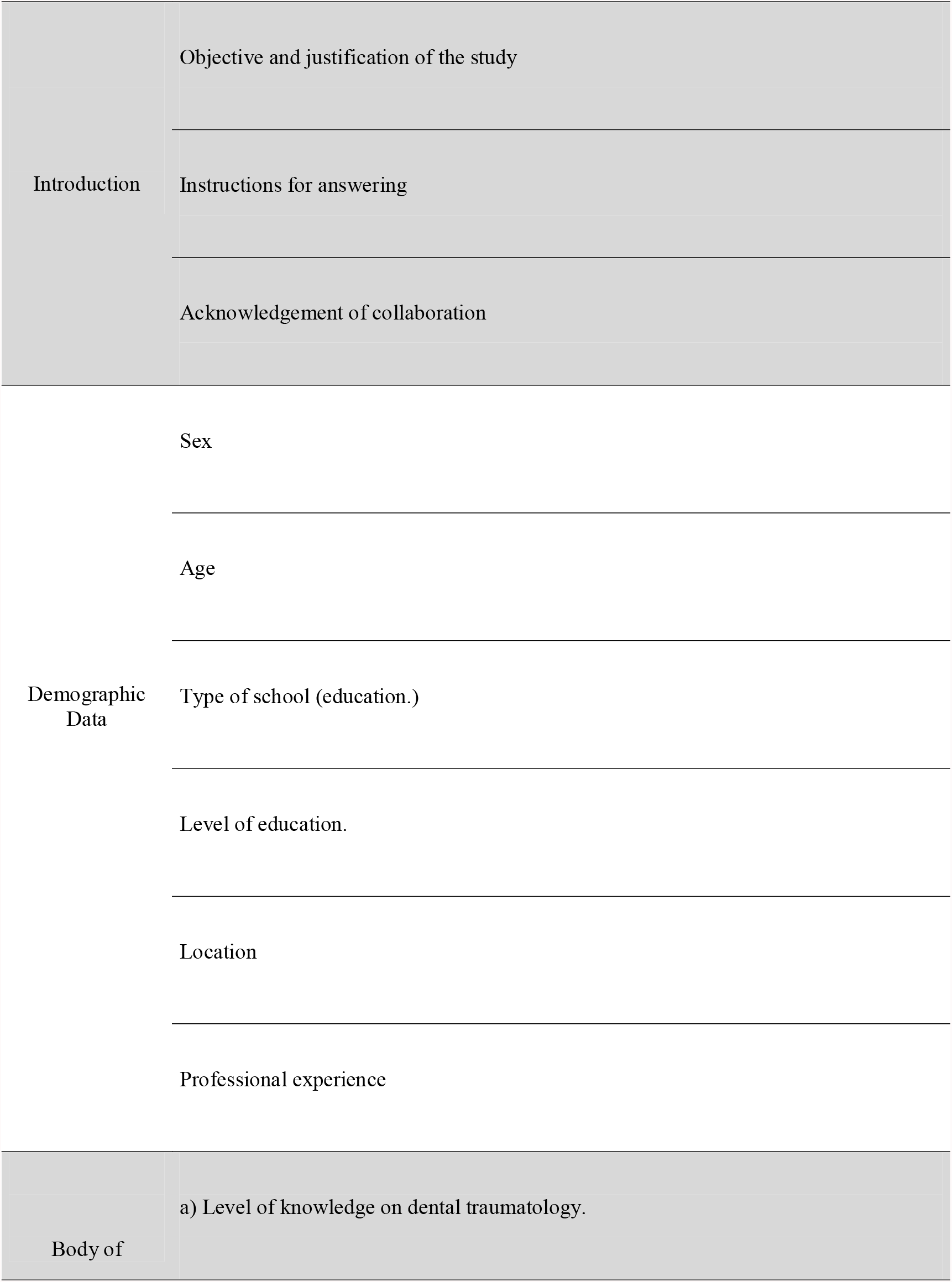
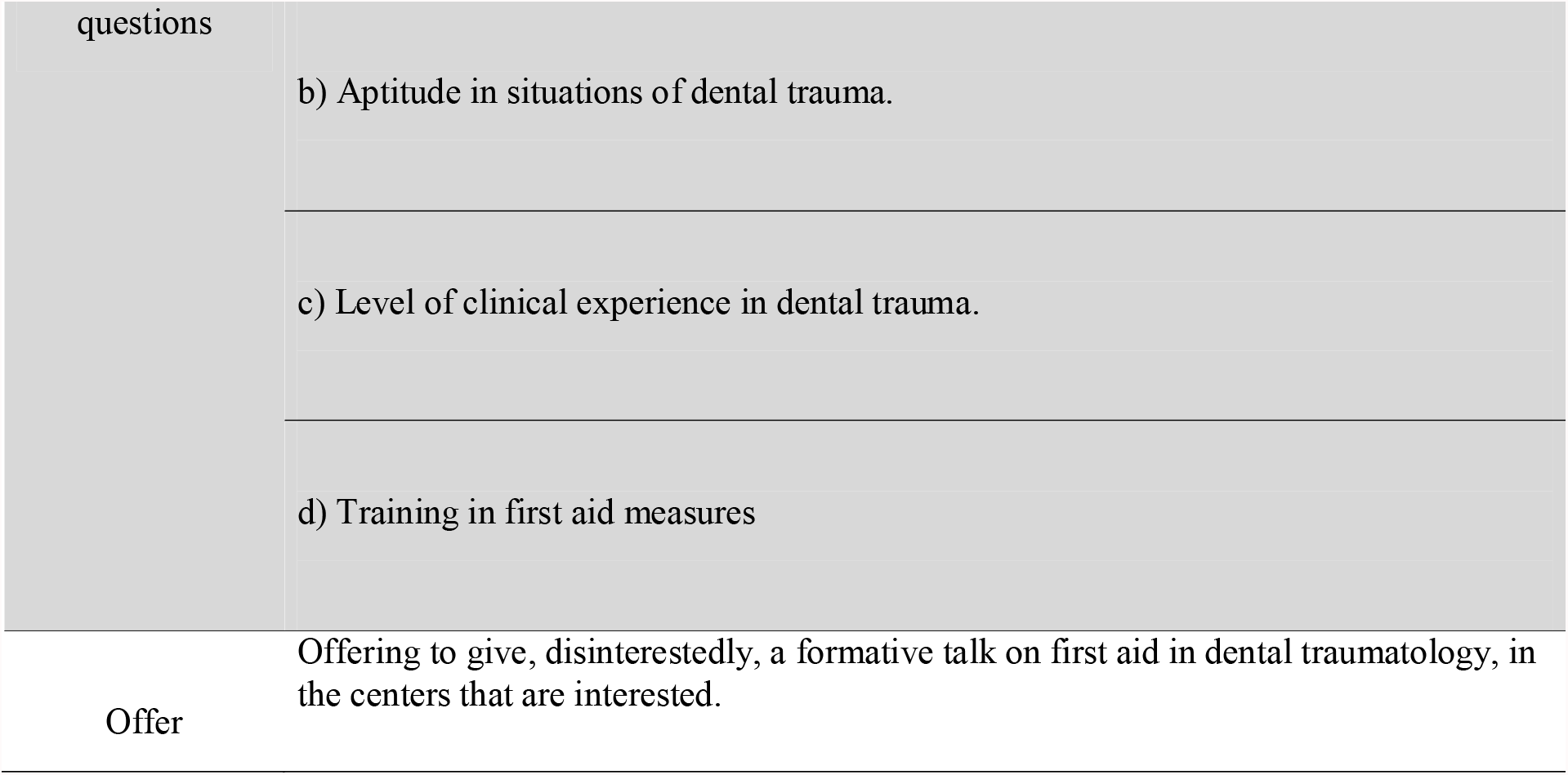
Structure (groups) and dimensions of the questionnaire on knowledge and aptitude of early childhood, primary and/or secondary education teachers referred to first aid measures in dental trauma.

The set of questions addressed 19 items organized into four dimensions:

a. Evaluation of the level of knowledge: four items assessing the knowledge of the participant referred to general concepts in dental trauma.
b. Evaluation of aptitude in relation to situations of dental trauma: eight items assessing the aptitude of the participant for providing care in the case of a pupil that has suffered tooth trauma.
c. Experience in dental trauma: two items exploring whether the participant has personally suffered dental trauma or helped someone else who has suffered dental trauma.
d. Training in first aid measures: five items assessing whether the participant has received training in first aid measures.

Scoring of the questionnaire Only the first two dimensions of the set of questions were scored, based on a scale of 0 to 1 points (0 points = incorrect answer and 1 = correct answer).

a. General knowledge of dental trauma: four multiple alternative closed response items, with only one correct answer.
b. Aptitude in relation to situations of dental trauma: eight multiple alternative closed response items, with only one correct answer. Since both dimensions comprised 4 and 8 items, respectively, the possible questionnaire scores ranged from 0-12 points. In order to categorize the quantitative variable “Questionnaire score” and transform it into an ordinal qualitative variable, we created the variable “Level of knowledge and aptitude”, with the definition of three levels. (table 4)

### Validation of the questionnaire

A pilot questionnaire was initially designed, comprising 30 questions and with a structure slightly different from that of the current final questionnaire. The pilot questionnaire was examined by 5 experts in the field, in order to assess its clarity and the pertinence of the questions. Their comments also contributed to define the wording and sequence of the questions in order to make the questionnaire as effective and efficient as possible. Application of the questionnaire The COVID-19 pandemic made it necessary to resort to online technologies (WhatsApp and social networks such as Facebook and Instagram) for distribution and application of the questionnaire, since population lockdown at the time of the study precluded contact through other means. The education and sports authorities of Andalusia provided the contact information corresponding to all the public centers in the province of Seville, thus allowing us to send e-mails to 670 out of a total of 799 centers, excluding those that failed to meet the study requirements. Likewise, the directors of the centers were contacted by mobile phone whenever possible.

Statistical analysis The data obtained from the questionnaire designed using the tool Google Forms were entered on MS Excel spreadsheets for subsequent analysis, with the exclusion of 16 questionnaires.

The qualitative data were coded and transformed into quantitative information for processing with the SPSS statistical package. A descriptive analysis was performed, with the generation of frequency tables and the calculation of measures of central tendency (mean and median), the verification of normal data distribution, the generation of cross-tables and calculation of Cramer’s coefficient of association to determine statistically significant correlations.

## RESULTS

The total sample size consisted of 442 participants representing 75 municipalities of the 106 found in the province of Seville. Table 3 shows the distribution of the sample according to gender and age, location, type of center, teaching level and professional experience.

**Table 3.** Distribution of the sample according to gender and age, location, type of center, teaching level and professional experience.

| Sex |  | Age |  | Locality |  |
| --- | --- | --- | --- | --- | --- |
| Male | 24,43% | 20-29 | 7,01% | Sevilla capital city | 41,85% |
| Female | 75,57% | 30-39 | 25,34% | Rest of Province | 58,15% |
|  |  | 40-49 | 38,91% |  |  |
|  |  | 50-59 | 24,21% |  |  |
|  |  | 60-65 | 4,53% |  |  |
| Center Ownership |  | Level of Education |  | Professional Experience |  |
| Concerted | 18,10% | Early Childhood 0 - 6 | 26,70% | < 1 year | 1,58% |
| Private | 5,43% | Primary 6 - 12 | 43,66% | 1-10 years | 26,92% |
| Public | 76,47% | Secondary 12 - 18 | 29,64% | 11-20 years | 40,05% |
|  |  |  |  | 21-30 years | 15,84% |

Of the 442 participants in the survey, 88.5% yielded a score of between 0-5 (out of 12 possible points), reflecting a low level of knowledge and aptitude in relation to first aid measures in dental trauma. Only 11.5% of the teachers scored between 6-9 points, corresponding to medium level of knowledge and aptitude. None of the participants scored above 9 points (high level of knowledge and aptitude) (Table 4).

**Table 4.**
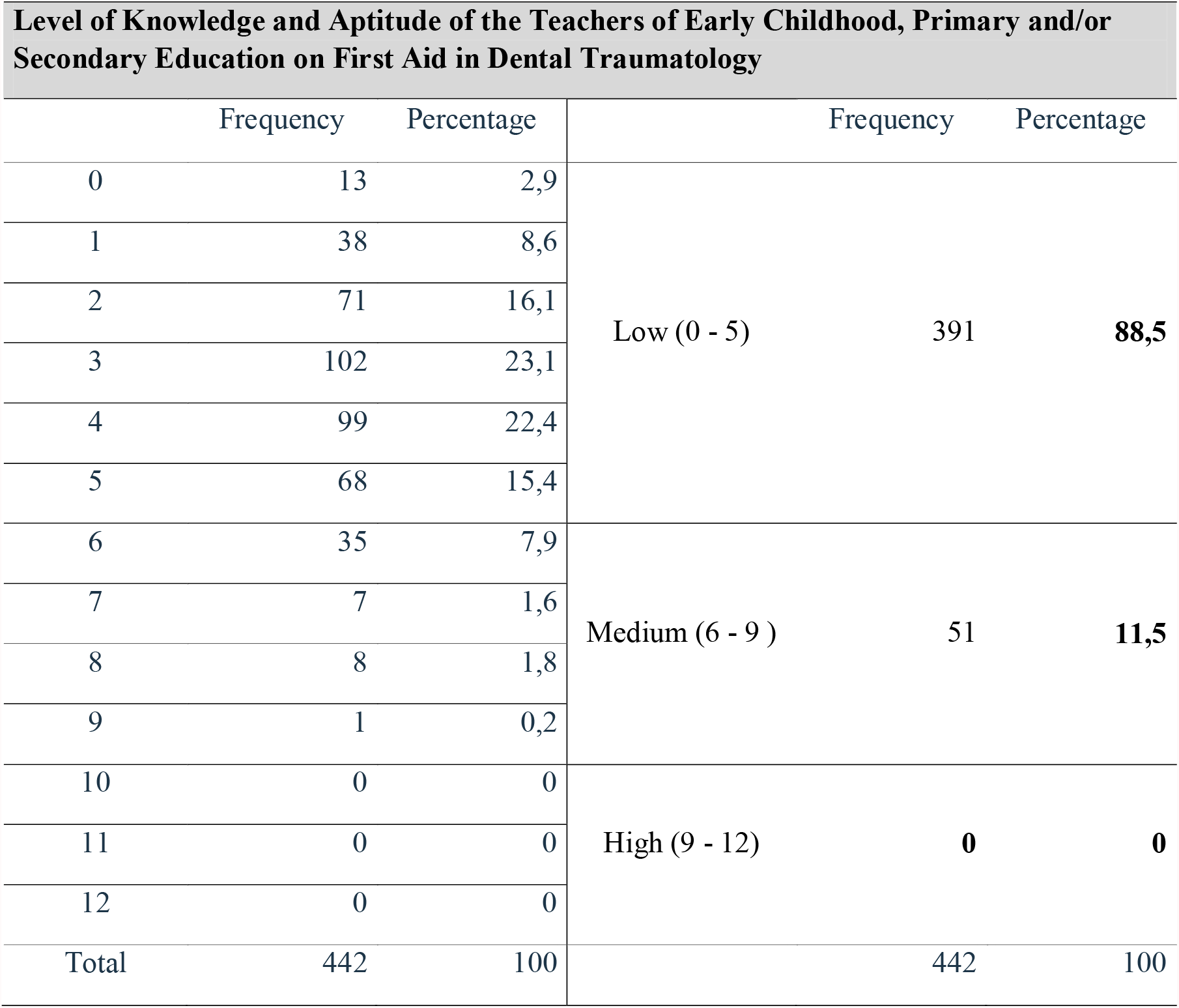
Level of knowledge and aptitude.

The participants scored better in the first dimension (knowledge) than in the second (aptitude). Specifically, in relation to the dimension “Level of knowledge”, 52.3% obtained medium level scores, and 8 even obtained high scores. In contrast, a full 90.7% produced low scores for the dimension “Aptitude”.

Tables 6, 7 and 8 respectively evidence the following: - Only 3.8% of the participants reported feeling able to reimplant an avulsed tooth. - Only 5.4% of the participants would transport the avulsed tooth in a container with milk. Of note is the observation that 70.1% would use toilet paper or a handkerchief. - Only 14% of the participants would go to the pediatric dentist; 39.8% would report to the primary care center; and 36.2% would go to the hospital emergency room.

**Table 5.**
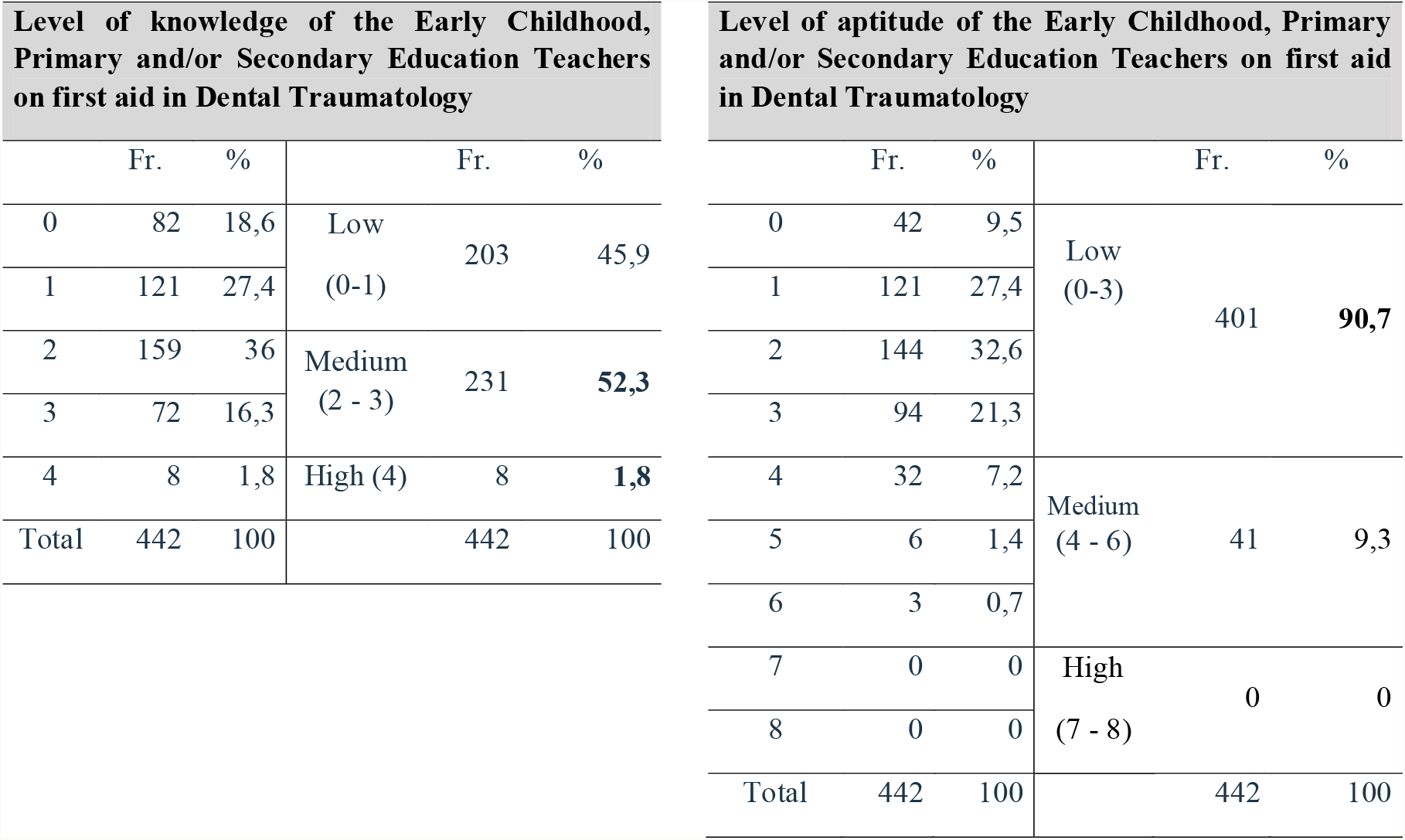
Level of knowledge versus level of aptitude in relation to first aid measures in dental trauma.

**Table 6.** Tooth avulsion reimplantation.

| **In case you find the permanent tooth, do you feel able to reimplant it?** |  |  |
| --- | --- | --- |
|  | Frequency | Percentage |
| I would look for health personnel from the school to do it | 32 | 7,2 |
| Don't know/ No answer | 14 | 3,2 |
| No, never | 372 | **84,2** |
| **Yes** | 17 | **3,8** |
| Only sometimes | 7 | 1,6 |
| Total | 442 | 100 |

**Table 7.** Storage medium and transport.

| **If you have the tooth, how would you transport it?** |  |  |
| --- | --- | --- |
|  | Frequency | Percentage |
| In the child's mouth | 11 | 2,5 |
| In toilet paper or a clean handkerchief | 310 | **70,1** |
| In a container of water | 20 | 4,5 |
| **In a container with milk** | 24 | **5,4** |
| In a container with saline solution | 77 | 17,4 |
| Total | 442 | 100 |

**Table 8.** Where to seek professional help.

| **If a student had a tooth broken or knocked out because of a severe blow to the mouth, where is the first place you would seek help?** |  |  |
| --- | --- | --- |
|  | Frequency | Percentage |
| Health Center or Primary Care | 176 | **39,8** |
| Dentist | 27 | 6,1 |
| **Paediatric Dentist** | 62 | **14** |
| Don't Know / No Answer | 17 | 3,8 |
| Emergency Room | 160 | **36,2** |
| Total | 442 | 100 |

**Table 9.** Level of knowledge and aptitude according to age.

|  |  |  | Age |  |  |  |  | Total |
| --- | --- | --- | --- | --- | --- | --- | --- | --- |
|  |  |  | 20-29<br>years | 30-39<br>years | 40-49<br>years | 50-59<br>years | 60-65<br>years |  |
| Level of<br>Knowledge<br>and Skills | Low | Count | 29 | 101 | 151 | 94 | 16 | 391 |
|  |  | %<br>within<br>age | 93,50% | 90,20% | 87,80% | 87,90% | 80,00% | 88,50% |
|  | Medium | Count | 2 | 11 | 21 | 13 | 4 | 51 |
|  |  | %<br>within<br>age | 6,50% | 9,80% | 12,20% | 12,10% | 20,00% | 11,50% |
| Total |  | Count | 31 | 112 | 172 | 107 | 20 | 442 |
|  |  | %<br>within<br>age | 100,00% | 100,00% | 100,00% | 100,00% | 100,00% | 100,00% |

**Table 10.** Level of knowledge and aptitude according to training received in relation to first aid measures in dental trauma.

|  |  |  | Training First Aid |  | Total |
| --- | --- | --- | --- | --- | --- |
|  |  |  | No | Sí |  |
| Level of Knowledge and Aptitude | Low | Count | 386 | 5 | 391 |
|  |  | % | 89,10% | **55,60%** | 88,50% |
|  | Medium | Count | 47 | 4 | 51 |
|  |  | % | **10,90%** | **44,40%** | 11,50% |
| Total |  | Count | 433 | 9 | 442 |
|  |  | % | 100,00% | 100,00% | 100,00% |

On the other hand, as the age of the participants increased, the percentage of those showing low knowledge and aptitude scores was seen to decrease (from 93.5% to 80%). Likewise, with increasing age the percentage of participants yielding médium scores was seen to rise (from 6.5% in those under 29 years of age to 20% in those over age 60).

The recorded Cramer’s coefficient of association of over 0.6 indicated a strong and statistically significant correlation between “Level of knowledge and aptitude” and the variable “Age” of the participant.

With regard to the training received, almost half (44.4%) of the teachers who had received training in first aid measures in dental trauma showed medium level of knowledge and aptitude. In contrast, this percentage dropped to only 10.9% among those teachers who had received no such training. Although Cramer’s coefficient of association indicated no statistically significant correlation between the variables, the analysis of the table appeared to indicate otherwise, since the proportion of teachers with medium level scores was higher in the group that received training than in the group without training.

## DISCUSSION

The clinical practice guides of the International Association for Dental Traumatology (IADT) confirm that the prognosis in cases of dental trauma is directly influenced by the time elapsed from injury to the start of adequate treatment [Diangelis et al, 2017 and Malmgrem et al, 2017.] There is scientific evidence and general agreement in dental traumatology that in certain clinical situations such as tooth avulsion, correct intervention at the site of the accident has an impact upon the prognosis [Andersson et al, 2017 and Blakytni et al, 2001.] Although many countries in Europe have implemented prevention and immediate intervention campaigns [Caglar et al 2005 and McIntyre et al, 2008], no such initiatives in oral health are found in Spain, designed to instruct and promote adequate intervention on the part of the population or teaching professionals in situations of dental trauma.

Taking into account that certain age groups (particularly the pediatric population) are more likely to suffer such injuries, it is advisable for those in charge of children (such as early childhood and primary and/or secondary education teachers) to be familiarized with a first aid intervention protocol in dental trauma. In our study, the teachers were not sufficiently trained in this area, as evidenced in 97.7% of the participants in the survey. This may be explained by the fact that a full 98% of the teachers claimed to have received no information about first aid measures in dental trauma through any means.

Studies similar to our own report somewhat more positive results in this regard, with an incidence of subjects able to deal with pediatric dental trauma (PDT) of under 25% (range 12-23.5%) [Tewari et al., 2020. Niviethitha et al., 2018. Tzimpoulas et al., 2020. Panwar et al. 2018. Alsadhan et al,. 2018.] Possibly as a consequence of the above, 88.5% of the participants in our survey presented a deficient level of knowledge and aptitude in relation to first aid measures in dental trauma.

A total of 15.6% of the participants expressed an interest in receiving training in this area. This low figure may be due to the fact that school campaigns usually focus on the prevention of caries, and to the widespread lack of knowledge of the frequency and sequelae of dental traumatisms.

In contrast to our results, other similar studies have reported a wish to receive more information about PDT on the part of most teachers [Tewari et al., 2020.] One of the factors that typically influence knowledge and aptitude in relation to certain subjects is gender. However, gender was not found to be relevant in our case, since very similar percentages were recorded for both men and women: 89.8% and 10.2% of the women showed low and medium scores, respectively, versus 84.3% and 15.7% of the men.

The differences between the two genders were not statistically significant. The teaching staff requirements vary depending on the type of center. However, our results show that regardless of the type of center (public, private or concerted), none have dedicated resources to ensure that their teaching staff receive basic training in this area. We observed no statistically significant relationship between the knowledge or aptitude of the teachers and the public or private nature of the center.

In contrast, a statistically significant relationship was observed between the level of knowledge and aptitude and the fact of having received training in dental first aid measures. Of the 9 teachers (2% of the sample) that had received training in this area, 44.4% (i.e., nearly half) showed a medium level of knowledge and aptitude. In comparison, of the 433 teachers (98% of the sample) that had received no such training, only 10.9% presented a medium level of knowledge and aptitude.

The aforementioned 2% of participants with training is much lower than in the rest of the studies found in our review of the literature. In effect, 8 studies reported first aid training in dental trauma in less than 75% of the participants [Tewari et al., 2020], while 20 studies reported figures of under 50%. [Niviethitha et al., 2018. Tzimpoulas et al., 2020. Panwar et al. 2018. Alsadhan et al,. 2018. Marcano-Caldera et al. 2018. Hassan et al., 2018a. Hassan et al. 2018b. Traebert, 2009.]

In reply to the question “If the tooth is broken, can the broken part be joined again?”, only 7% were aware that joining is possible in the case of a permanent tooth. Ten studies in other countries have found that 50% of the teachers are aware of the importance of finding the broken tooth fragment [Tewari et al., 2020. Panwar et al,. 2018.Traebert, 2009.]

In reply to the question “If you find the permanent tooth, would you feel able to reimplant it?, a full 84.2% of the participants in our survey stated that they would not be able. While the possession of knowledge referred to performing immediate reimplantation of a permanent tooth (recorded in 5 out of 6 studies in a meta-analysis published in 2020 [Tewari et al., 2020] or no immediate reimplantation of a temporary tooth (recorded in 7 out of 10 studies in the mentioned meta-analysis [Tewari et al., 2020] is important, it is also necessary to feel able to do so in a critical moment [McIntyre et al., 2008.]

With regard to the storage and transport medium of the tooth, only 5.4% were aware that milk is the ideal medium. Only 25% of the teachers in other studies [Panwar et al. 2018. Alsadhan et al,. 2018. Marcano-Caldera et al. 2018. Hassan et al., 2018a. Hassan et al. 2018b. Traebert, 2009] were aware of this fact, or cited the saliva of the affected child as the indicated medium. Of note is the observation that 70.1% of the participants in our survey would use toilet paper or a handkerchief, which would cause the death of the periodontal ligament cells and thus result in irreversible damage. In the present study, only 15% of the participants would seek specialized help from a pediatric dentist.

In contrast, in our review of the literature, over 70% of the teachers would report to a doctor or specialist for adequate management [Blakytny et al., 2001. Tewari et al., 2020.] These data evidence a clear lack of preparation in these aspects compared with other countries, despite the fact that the required knowledge is easy to assimilate for teaching professionals. Only 3.8% would reimplant the tooth, despite the fact that viability of the latter increases if the period dry outside the mouth is less than 20 minutes. This means that viability could be facilitated provided reimplantation is made at the site of the accident.

Ten out of 17 studies conducted in other geographical settings have reported knowledge of the importance of the time of reimplantation [Tewari et al., 2020.]

The time factor is crucial in dental trauma, and in this regard 54.1% of the participants considered it desirable to seek help in under 30 minutes. The mistake of reporting first to a primary care center (39.8% of the participants in the survey) or to the hospital emergency room (36.2%) rather than to a dentist (6.1%) or more specifically to a pediatric dentist (14%) could be avoided through adequate training on how to deal with a dental emergency. With acquisition of the required knowledge, the observed figures could be expected to improve considerably, since they are a consequence of a serious lack of information. In any case, we found that with advancing age, the incidence of a low level of knowledge and aptitude decreased among the teachers (from 93.5% to 80%), while the incidence of a moderate level of knowledge and aptitude increased (from 6.5% in those under 29 years of age to 20% in those over age 60) – though Cramer’s coefficient of association did not find this correlation to be statistically significant. The present study is limited to the province of Seville (Spain), but this is not a local problem, and other studies have reported very similar data.

In a study conducted in 2009 in Brazil, only 2.2% of the teachers were found to have good levels of knowledge [Traebert, 2009.] Another study carried out in the United States in turn found that 44% of the surveyed teachers did not feel able to reimplant a tooth, and that 28% did not know how to do so [McIntyre et al., 2008.]

A European study in 2005 found that 74.3% of the surveyed teachers admitted having no knowledge about dental trauma [Caglar et al., 2005.] Lastly, a study carried out in the United Kingdom in 2001 found that 66.1% of the study subjects had received no information or advice about dealing with dental trauma [Blakytny et al., 2001.]

## CONCLUSIONS

The early childhood and primary and/or secondary education teachers in the province of Seville (Spain) present marked deficiencies in their level of knowledge and aptitude referred to the adoption of first aid measures in dental trauma among their pupils. Institutional campaigns are needed to improve the level of knowledge in this field among the teachers of the province.

## Data Availability

All data produced in the present study are available upon reasonable request to the authors

## Ethical approval

All procedures in this study involving human subjects were carried out in compliance with the ethical standards of the Declaration of Helsinki (1964 and subsequent amendments) or comparable standards, and the study protocol was approved by the Ethics Committee of the University of Seville (Spain).

## Informed consent

Informed consent was obtained individually from the participants in the study

## ACKNOWLEDGEMENTS

We’d like to thank all the school teachers who made possible this study.

All the authors declare that the manuscript is not being considered for publication in another journal.

## Notes

### Competing Interest Statement

The authors have declared no competing interest.

### Funding Statement

This study did not receive any funding

### Author Declarations

Ethics committee of University of Seville gave ethical approval for this work

